# Assessing the community risk perception toward COVID-19 outbreak in South Korea: evidence from Google and NAVER relative search volume

**DOI:** 10.1101/2020.04.23.20077552

**Authors:** Atina Husnayain, Eunha Shim, Anis Fuad, Emily Chia-Yu Su

## Abstract

This study aimed to explore the patterns of community health risk perception of coronavirus disease 2019 (COVID-19) in South Korea using Internet search data. Google and NAVER relative search volume data were collected using COVID-19-related terms in Korean language. Online queries were compared with the number of new COVID-19 cases and tests. Time series trends and Spearman’s rank correlation coefficients showed that the number of COVID-19-related queries in South Korea increased during the local and international events; higher in women, certain age groups; and higher in affected areas, which represented the community health risk perception. Greater correlations were found in mobile searches compared to that of desktop searches, indicating the changing behavior in searching health online information. The use of both Google and NAVER RSV to explore the patterns of community health risk perception could be beneficial for targeting risk communication in several perspectives including time, population characteristics, and location.

**Article Summary Line:** The use of both Google and NAVER RSV to explore the patterns of community health risk perception toward COVID-19 in South Korea could be beneficial for targeting risk communication in several perspectives including time, population characteristics, and location.

## Introduction

The World Health Organization (WHO) has already declared the coronavirus disease 2019 (COVID-19) outbreak as a pandemic since March 11, 2020 *(1)*. As of April 6, 2020, the disease has infected 1,210,956 individuals worldwide including 10,284 individuals in South Korea *(2)*. The first COVID-19 case in South Korea was confirmed on January 20, 2020 *(3)*. Slow upturns of disease transmission were reported before February 19, 2020; the huge local clusters observed in Daegu led to the increases in the number of new cases daily *(4)*. Numerous approaches have been conducted to prevent disease transmission along with coronavirus drive-through tests and social distancing *(5, 6)*. Coronavirus drive-through tests were identified as a safe and efficient screening approach, with each test taking approximately 10 minutes, and thus minimize the cross-infection among testees *(6)*. To date, the average number of daily new cases is now three times lower than those during the peak of the epidemic (from February 19 to March 15, 2020) *(3)*. Consequently, South Korea has been considered among the best-performing countries to tackle the pandemic.

On the contrary, adequate risk communication could also help minimize the impact of disease spread *(7)*. Thus, in the pandemic period, the WHO suggests regular risk communication by updating any changes in the status of pandemic to the public and stakeholders *(8)*. This action might be challenging since proper risk communication needs a robust understanding of risk perception which could identify what knowledge the public needs *(7)*. However, studies exploring risk perception were often conducted using survey methods or content analysis *(7, 9-11)*, which requires huge resources and takes longer time especially when investigating an emerging disease. This approach might be less affordable since the health system will be overburdened with the surge of healthcare utilizations, thus causing more barriers for assessing the community health risk perception.

Therefore, this study aimed to explore the patterns of community health risk perception towards COVID-19 in South Korea using Internet search data. This novel approach is potentially used so that the Internet query data could be provided more easily, in a timely manner, and in a cost-effective way compared with the survey method *(12)* and also potentially capture anomalous patterns in real time *(13)*. In this analysis, we utilized Google and NAVER relative search volume (RSV) to represent the online queries from the biggest search engine and Korean local search engine. This study explored the patterns of public health risk perception towards the ongoing outbreak in several different perspectives including time, population characteristics, and location as used in epidemiological studies. Future studies are warranted in order to define the best lagged period in performing effective risk communication in early stage of disease outbreak.

## Methods

### Datasets

Numbers of new COVID-19 cases and coronavirus tests performed on a daily basis were collected from the South Korea open access data set from Kaggle by Joong Kun Lee and colleagues in collaboration with the Korea Centre for Disease Control and Prevention (KCDC) from January 20 to March 22, 2020 *(3)*. We used the Time.csv dataset to retrieve the number of new daily COVID-19 cases and tests on a daily basis and TimeProvince.csv dataset to collect the cumulative coronavirus cases by region. Those datasets covered all cities in South Korea. We used all data provided, including those during the observation period. By contrast, Internet search data related to COVID-19 were retrieved from Google Trends (https://trends.google.com/) and NAVER websites (https://datalab.naver.com/) in the same geolocation. The information searched were collected six weeks earlier from December 5, 2019, to explore the patterns prior to the occurrence of the first COVID-19 case in South Korea. Google and NAVER RSV data were collected using COVID-19-related terms including coronavirus (코로나 바이러스), coronavirus test (코로나 바이러스 테스트), MERS (메르 스), facemask (마스크), and social distancing (사회적 거리두기) in Korean language and retrieved according to time, gender, age groups, type of devices, and location.

### Analysis

The data were analyzed in a single graphical form to explore the trends in the new COVID-19 cases, number of tests, and Internet searches on a daily basis. Time-lagged correlation calculated by Spearman’s rank correlation coefficients were employed to assess whether correlations between new COVID-19 cases, Google, and NAVER RSV were affected by time within 3 days of lagged and lead period. The statistical analysis was performed using STATA13, and strong correlations were defined as correlation coefficients greater than 0.7.Moreover, multilayer maps created using Tableau Public were generated to define the distributions of new COVID-19 cases and Internet searches.

## Results

Community health risk perception captured by Google and NAVER RSV were divided into several parts, including patterns by time, population characteristics, and location:

### Trends in new COVID-19 cases, number of tests, and Internet searches on a daily basis

South Korea reported the first case of COVID-19 on January 20, 2020, as shown in Figure 1 with three peaks of disease transmissions. The first peak occurred until February 18, 2020. The average new cases increased to 311 and dramatically decreased to 110 cases per day since March 16, 2020. As of March 22, 2020, South Korea reported 8,897 cases of COVID-19. On the contrary, an immense number of tests were also performed during the outbreak. South Korea has performed 5,266 tests per day on average from January 20 to March 22, 2020, and 466,804 tests in total as of April 6, 2020, recording South Korea as a country with the third highest number of tests performed.

**Figure 1.**
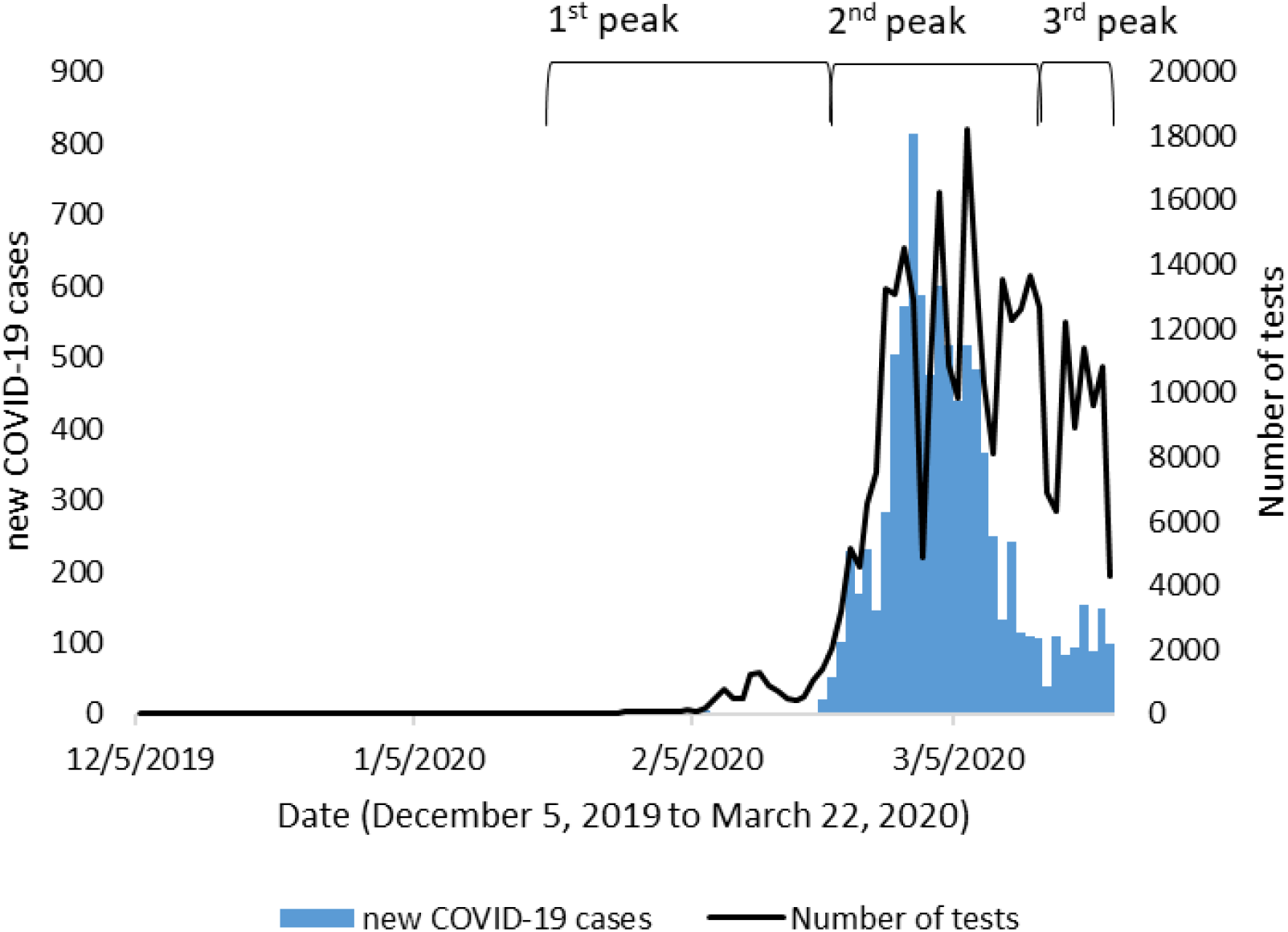
Time series of new COVID-19 cases and number of tests in South Korea.

During the outbreak, trends of information searches for coronavirus (코로나 바이러스) captured by Google and NAVER were similar as shown in Figure 2(A). Three huge peaks of Internet searches were observed in the second and fifth week of January and in the fourth week of February 2020. Coronavirus-related searches remained high for several days since the first COVID-19 case was reported in Wuhan on December 12, 2019 along with MERS (메르 스)-related queries which also elevated in last two peaks. However, massive surges of information searches occurred along with the identification of the first COVID-19 case in South Korea on January 20 and the WHO’s declaration of the Public Health Emergency of International Concern (PHEIC) on January 30, 2020. Compared with the daily data on new COVID-19 cases, information searches provided by Google Trends and NAVER peaked six to seven days earlier. The third peak of coronavirus searches possibly correspond to the immense increase in the number of new COVID-19 cases due to local transmission. Searches gradually decreased even after the outbreak was declared as a pandemic by the WHO on March 11, 2020 *(1)*.

**Figure 2.**
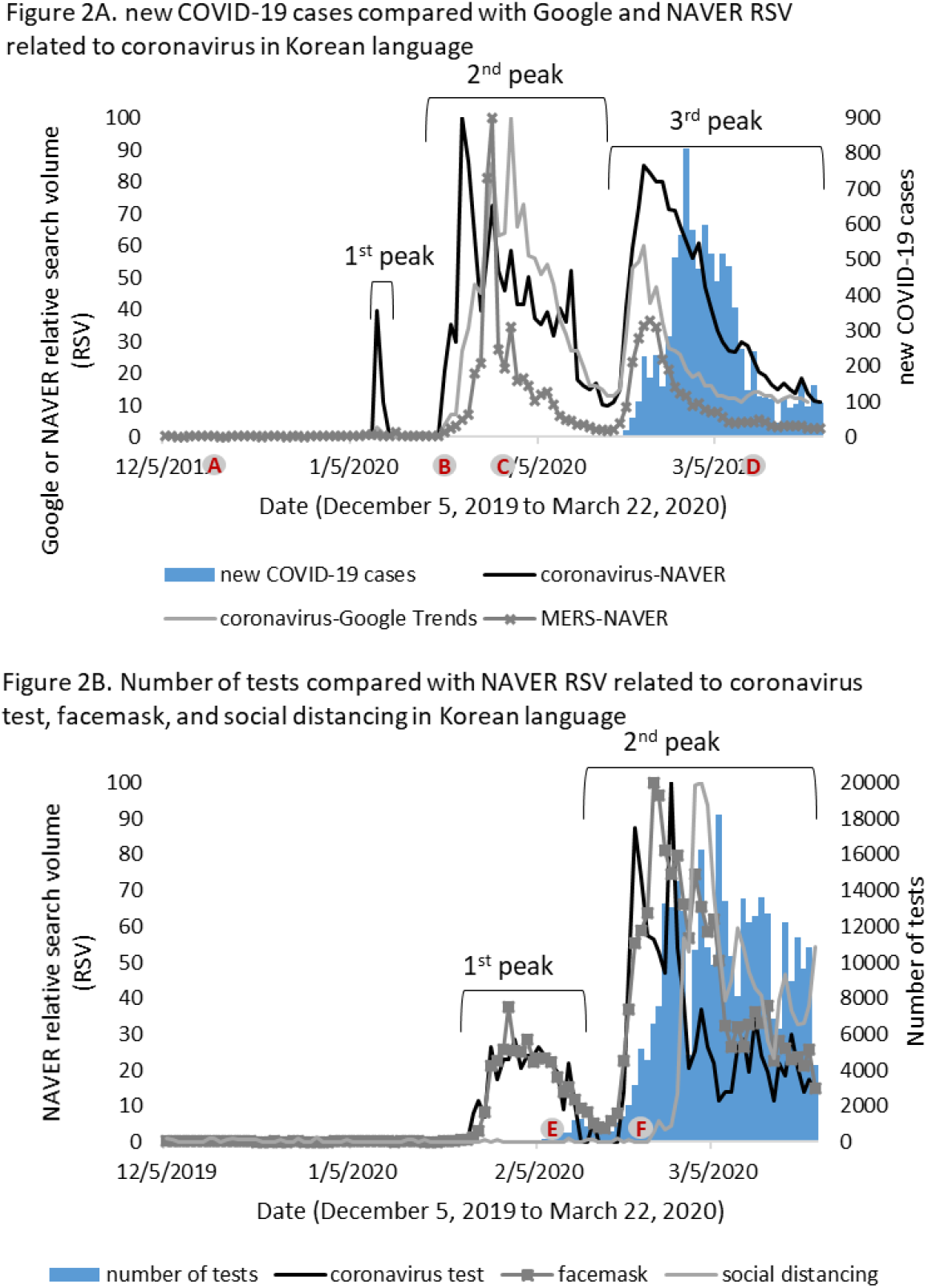
Time series of new COVID-19 cases, number of tests, Google, and NAVER RSV related to coronavirus in South Korea.

Furthermore, coronavirus test-related searches (코로나 바이러스 테스트) were not captured in Google Trends; hence, Figure 2(B) only showed NAVER RSV related to coronavirus tests, facemask, and social distancing. Increases in the Internet searches were observed weeks after the COVID-19 cases were reported and before coronavirus test kit was approved on February 7, 2020 *(14)*. The second wave of information searches was found in the third week of February 2020, which might be caused by the increase in the number of new COVID-19 cases and the implementation of coronavirus drive-through tests on February 23, 2020 *(6)*. However, patterns of coronavirus test-related searches seemed more similar to trends of new COVID-19 cases compared with the daily numbers of tests.

Similar patterns to the online queries on coronavirus test were also identified for facemasks (마스크). In the role of personal protective measures, facemask-related queries were elevated in the same period when people started to search for coronavirus tests and facemask shortage in early February *(15)*, and gradually declined in late of February as the regular supply of facemask has been provided government *(16)*. Besides, the massive increase of locally acquired cases also induced huge internet searches related to social distancing (사회적 거리두기) as one of the preventive approaches. Those searches reached their peaks as a widespread campaign for social distancing in the first week of March 2020 in South Korea *(5)*.

### Time-lagged correlations between new COVID-19 cases and Internet searches in different gender and age groups

As shown in Table 1, results demonstrated a moderate correlation (0.640) between new COVID-19 cases and Google RSV related to coronavirus in lag −3. On the contrary, the high correlations (0.718) of coronavirus information searches counted for both men and women in lag −3 showed no differences for NAVER RSV. However, the correlations varied across different age groups and lagged periods. High correlations were observed in lag −3 for overall ages (0.729), and those aged ≤18 years (0.821), 19−24 years (0.784), 25−29 years (0.726), 50−54 years (0.706), and ≥50 years (0.725). Meanwhile the lowest correlation was found at the age group of 35−39 years old (0.622). The ≤18 year and 19−24 year age groups for NAVER RSV have high correlations in almost all lagged and lead periods. Moreover, the strength of correlations was decreased in the lead period or a few days after the number of new COVID-19 cases increased, either for Google or NAVER RSV. Compared to NAVER RSV, Google RSV for coronavirus has lower correlations with new COVID-19 cases.

**Table 1.** Time-lagged correlation coefficients between new COVID-19 cases, Google, and NAVER RSV related to coronavirus and coronavirus test in South Korea.

| coronavirus (코로나 바이러스) |  |  |  |  |  |  |  |  |  |  |  |  |  |
| --- | --- | --- | --- | --- | --- | --- | --- | --- | --- | --- | --- | --- | --- |
| Day- | Google Trends | NAVER |  |  |  |  |  |  |  |  |  |  |  |
|  |  | Gender |  | Age groups (years old) |  |  |  |  |  |  |  |  |  |
|  |  | Male | Female | Overall | ≤18 | 19-24 | 25-29 | 30-34 | 35-39 | 40-44 | 45-49 | 50-54 | ≥55 |
| -3 | **0.64** | **0.718** | **0.718** | **0.729** | **0.821** | **0.784** | **0.726** | **0.661** | **0.622** | **0.648** | **0.685** | **0.706** | **0.725** |
| -2 | 0.611 | 0.684 | 0.684 | 0.694 | 0.805 | 0.759 | 0.696 | 0.621 | 0.581 | 0.607 | 0.655 | 0.68 | 0.693 |
| -1 | 0.594 | 0.67 | 0.67 | 0.681 | 0.812 | 0.759 | 0.678 | 0.601 | 0.561 | 0.593 | 0.638 | 0.662 | 0.682 |
| 0 | 0.59 | 0.654 | 0.654 | 0.663 | 0.803 | 0.737 | 0.659 | 0.578 | 0.538 | 0.565 | 0.606 | 0.634 | 0.655 |
| 1 | 0.569 | 0.647 | 0.647 | 0.661 | 0.794 | 0.736 | 0.66 | 0.579 | 0.536 | 0.56 | 0.606 | 0.633 | 0.658 |
| 2 | 0.532 | 0.591 | 0.591 | 0.606 | 0.759 | 0.688 | 0.6 | 0.513 | 0.477 | 0.508 | 0.554 | 0.58 | 0.606 |
| 3 | 0.512 | 0.579 | 0.579 | 0.597 | 0.749 | 0.682 | 0.587 | 0.5 | 0.468 | 0.498 | 0.537 | 0.565 | 0.592 |
| coronavirus test (코로나 바이러스 테스트) |  |  |  |  |  |  |  |  |  |  |  |  |  |
| Day- | Google Trends | NAVER |  |  |  |  |  |  |  |  |  |  |  |
|  |  | Gender |  | Age groups (years old) |  |  |  |  |  |  |  |  |  |
|  |  | Male | Female | Overall | ≤18 | 19-24 | 25-29 | 30-34 | 35-39 | 40-44 | 45-49 | 50-54 | ≥55 |
| -3 | N/A | 0.739 | 0.769 | 0.77 | **0.595** | 0.681 | 0.654 | 0.701 | 0.734 | 0.696 | 0.624 | **0.612** | 0.441 |
| -2 | N/A | 0.769 | 0.79 | 0.797 | 0.505 | 0.65 | 0.687 | 0.752 | 0.786 | 0.692 | **0.673** | 0.581 | 0.445 |
| -1 | N/A | **0.795** | 0.799 | 0.824 | 0.5 | **0.725** | 0.645 | 0.775 | **0.826** | 0.704 | 0.63 | 0.532 | 0.434 |
| 0 | N/A | 0.778 | 0.799 | 0.812 | 0.542 | 0.72 | 0.653 | 0.746 | 0.783 | **0.755** | 0.559 | 0.551 | 0.358 |
| 1 | N/A | 0.775 | **0.823** | **0.828** | 0.508 | 0.682 | **0.688** | **0.786** | 0.814 | 0.718 | 0.586 | 0.557 | **0.45** |
| 2 | N/A | 0.756 | 0.802 | 0.805 | 0.549 | 0.62 | 0.623 | 0.774 | 0.762 | 0.731 | 0.586 | 0.537 | 0.433 |
| 3 | N/A | 0.744 | 0.763 | 0.781 | 0.465 | 0.572 | 0.606 | 0.694 | 0.756 | 0.633 | 0.633 | 0.518 | 0.424 |

Note: All correlations are statistically significant at a p-value of ≤0.05. Shade text: high correlation with r>0.7. Text in italic: the highest correlation.

Different patterns occurred for coronavirus test-related search. No correlation could be calculated for Google RSV due to the insufficient number of queries recorded. High correlations were found in lag −1 for men (0.795) and lead 1 for women (0.823) for NAVER RSV, as well as for all age groups in lead 1 (0.828). Moreover, weak to strong correlations were reported in different age groups. The 19–24-year age group has a high correlation (0.725) in lag −1 followed by the 30–34-year age group (0.786 in lead 1), 35–39-year age group (0.826 in lag −1), and 40– 44-year age group (0.755 in lag 0), respectively.

### Trends in online information searches based on the type of devices used for assessing Internet

Figure 3(A) showed the trends of online information searches for coronavirus and coronavirus tests in mobile devices and desktops. Mobile search queries for coronavirus were in-line with the desktop search during the first peak. However, mobile searches were higher either in the second or third peak of the outbreak. For coronavirus test-related searches, mobile searches seemed to be more frequent and stable than those of desktop searches, in all peaks.

**Figure 3.**
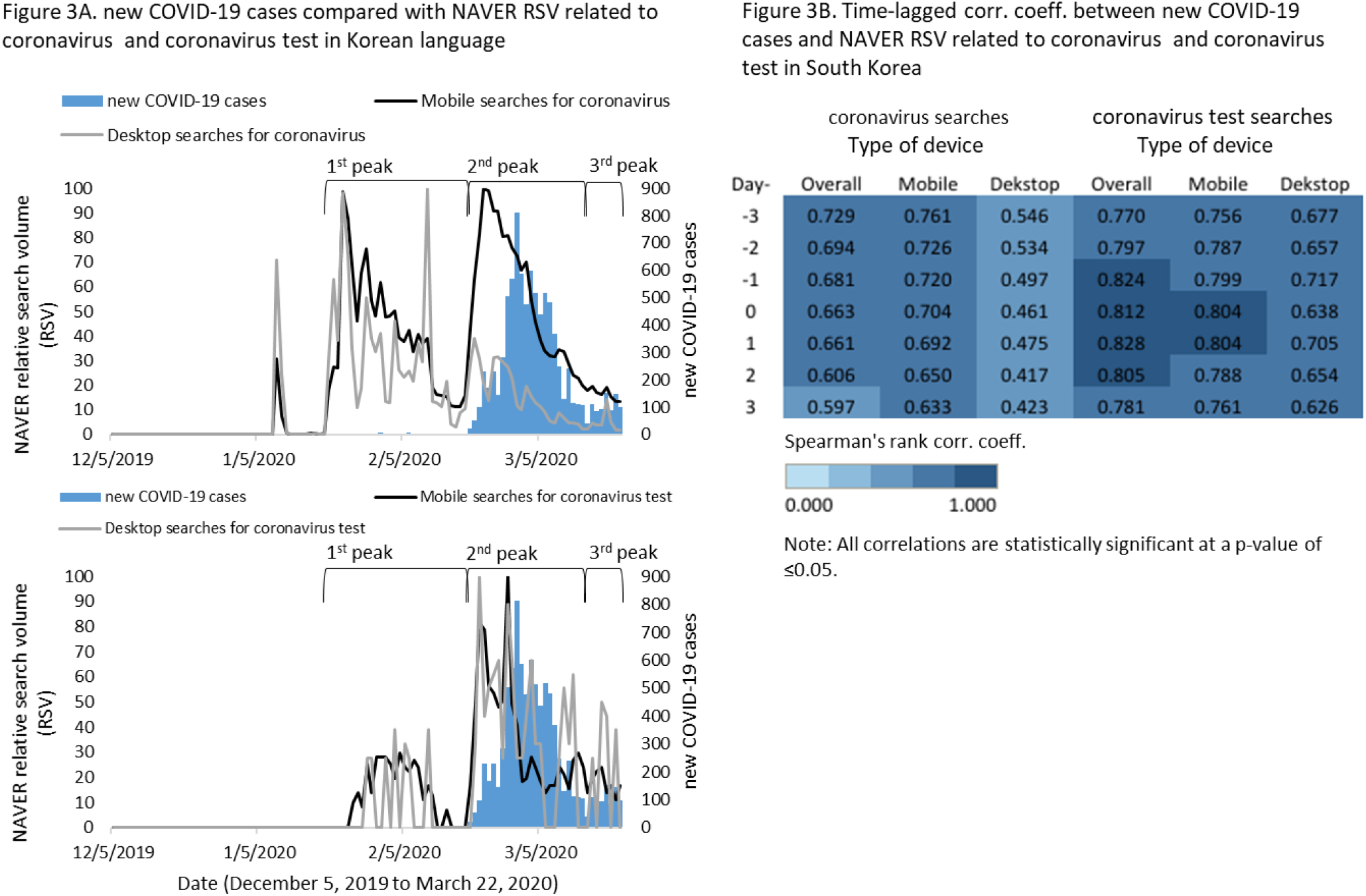
Time series and time-lagged correlation coefficients between new COVID-19 cases and NAVER RSV related to coronavirus for mobile and desktop searches in South Korea.

Spearman’s rank correlation coefficient in Figure 3(B) showed high correlations for overall dataset (mobile and desktop searches) of coronavirus searches in lag −3 (0.729), as well as mobile searches (0.761). Interestingly, mobile searches have stronger correlation coefficients in all lagged and lead periods than the overall searches. Yet, weak-to-moderate correlations (0.417–0.546) were observed for coronavirus-related searches through desktop devices. For coronavirus test online searches, high correlations (0.770–0.828) were reported in all lagged and lead days. Still, mobile searches were observed to have a higher correlation coefficient than desktop searches. The highest correlation found in lag 0 for mobile searches (0.804) and lag −1 for desktop searches (0.717).

### Distributions of new COVID-19 cases and Internet searches

Spatial distributions of new COVID-19 cases and Google RSV are illustrated in Figure 4. Results showed that 9 days before the confirmed cases were reported in South Korea, the numbers of Google RSV related to coronavirus captured in Gyeonggi-do, Seoul, and Incheon Province increased. Then, the aforementioned provinces reported COVID-19 confirmed cases. During the early weeks of disease transmission, COVID-19 spread in Seoul, Incheon, Gwangju, Gyeonggi-do, and Jeollabuk-do as shown in Figure 4(B-D). Similar patterns were also captured for Google RSV that seemed to be elevated along with those periods in the western part of South Korea where the confirmed cases were reported.

**Figure 4.**
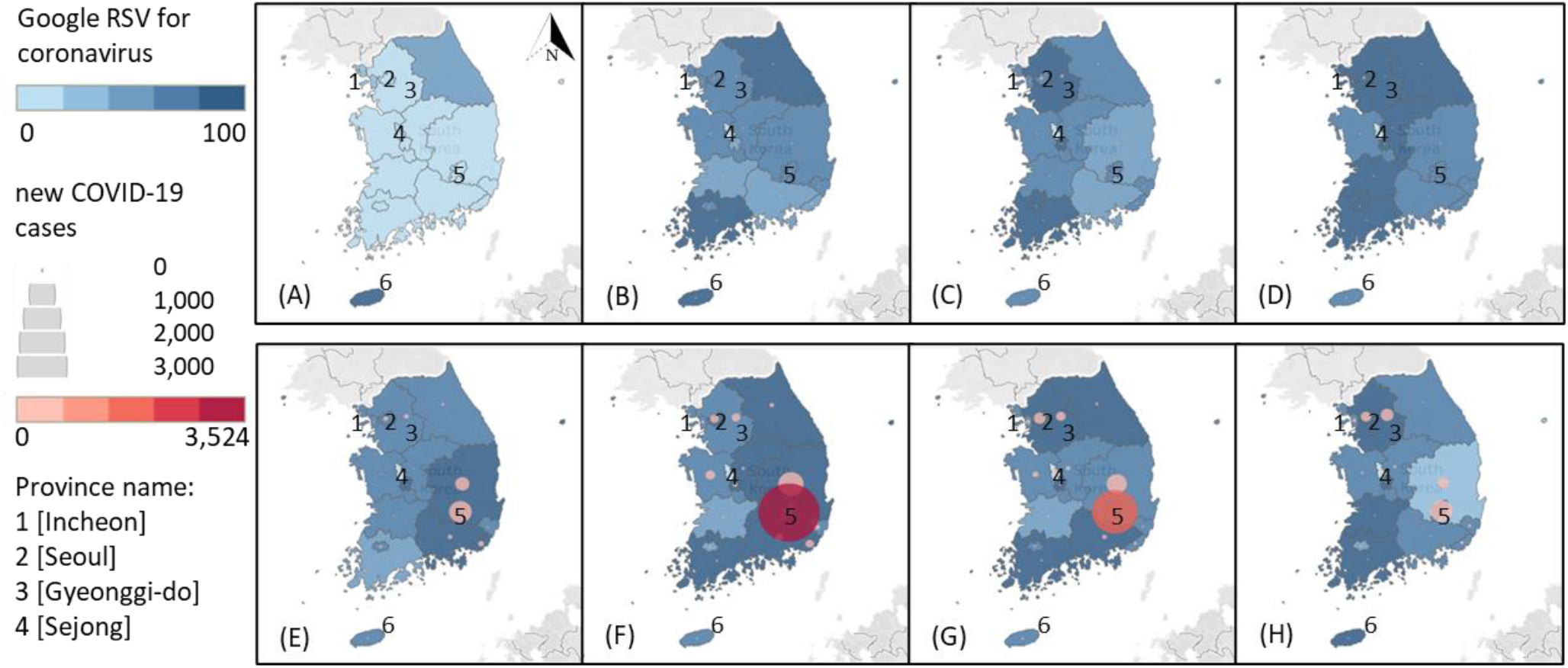
The numbers of new COVID-19 cases and Google RSV in South Korea: (A) January 11, 2020 to January 19; (B) January 20, 2020 to January 28, 2020; (C) January 29, 2020 to February 6, 2020; (D) February 7, 2020 to February 15 2020; (E) February 16, 2020 to February 24, 2020; (F) February 25, 2020 to March 4, 2020; (G) March 5, 2020 to March 13 2020; (H) March 14, 2020 to March 22 2020

Furthermore, a huge surge of new COVID-19 cases started on February 19, 2020, as shown in Figure 4(E). Google RSV gradually increased during those periods for the eastern part of South Korea including Daegu, the epicentre for local transmission, which contributed to 71.79% of the confirmed cases or 262.14 cases per 100,000 population *(17)*. Increases in online searches were observed in Gangwon-do, Gyeongsangbuk-do, Chungcheongbuk-do, Daejeon, Gyeongsangnam-do, and Jeollanam-do, the surrounding provinces of Daegu. The massive numbers of locally acquired cases occurred from February 25 to March 4, 2020 (Figure 4(F)) and quickly declined in the mid of March (Figure 4(G)). When number of new cases decreased, the number of Internet searches in the western part of South Korea started to increase, which showed an elevation in the number of COVID-19 cases in the later part of the study period (Figure 4(H)). Interestingly, Internet queries remained high (66-100 points) in Jeju, one of the most famous tourist destinations, during the period of observation; meanwhile, the incidence rate was only 0.60 per 100,000 population. By contrast, the incidence of COVID-19 in Sejong was 12 times higher (11.98 per 100,000 population) than that in Jeju. However, the number of search queries throughout the observation period was low.

## Discussion

Risk perception was defined as a person’ subjective judgments toward the likelihood of negative occurrences including diseases or illnesses *(18)*. In terms of disease outbreak, understanding the community health risk perception is urgently needed in the early phase of an outbreak particularly in the case of an emerging disease. This is because, in the initial period, there will be limited treatments, few numbers of resources, and delays in active interventions *(19)*. Therefore, exploring the perception of risk would be a necessary step in managing the risk of outbreak. Since a robust public risk perception assessment could help in divining effective risk communication, this step should be conducted immediately to reduce the impact of the COVID-19 outbreak. Consequently, it would be more affordable to conduct the community health risk perception assessment using Internet search data, since it could be provided more easily, in a timely manner, and in a cost-effective way compared with a survey method *(12)* and also potentially capture anomalous patterns in real-time *(13)*. With the widespread use of mobile devices and Internet, Internet search data could be more accurate in representing the community health risk perception *(20)* as information seeking intention is directly affected by risk perception *(9)*.

In this study, we found various correlations, which ranged from weak to strong correlations, among Google, NAVER RSV, new COVID-19 cases and number of tests. Previous studies also reported high correlations between Google and NAVER RSV compared with surveillance data *(12, 21)*. Therefore, immense searches of COVID-19-related information might represent the community health risk perception during local and international events. NAVER RSV, as a local search engine that obtained the second largest market share in South Korea, tends to be more sensitive against local issues such as coronavirus test as shown in Figure 2(B). A similar result has also been reported in a previous study which demonstrated that Baidu has better predictive performance for disease prediction than Google RSV *(21)*. These findings suggest that NAVER RSV could also potentially complement the use of Google RSV, which is excessively utilized in the field of surveillance and health risk perception assessment.

Patterns of community risk perception retrieved from information searches in this analysis were explained in different aspects: time, gender, age groups, type of devices used for accessing Internet, and spatial distributions. Patterns according to time showed that online queries related to COVID-19 increased during local events including the local transmission, approval of coronavirus test kits, implementation of coronavirus drive-through tests, facemask shortage, and widespread campaign for social distancing as well as during international events such as the announcement of PHEIC. Yet, South Korea was also one of the countries affected by the MERS epidemics *(22)*. That experience might also be one of the reasons for the increased searches for coronavirus even though cases have not yet been detected. These findings indicated that public health risk perception increased following both local and international crises. Hence, risk communication should be conducted promptly, considering that health risk perception might be change over time as the outbreak progresses.

Patterns by time also revealed the decreased number of Google and NAVER RSV in the middle of the epidemic curve, which might be caused by the extensive availability of online news and health expert reports during this period *(23)*. It might also be provoked by the decreased risk perception as the epidemic progressed *(7)*. Thus, utilizing Internet query data in analyzing community risk perception could be useful in the early stage of an outbreak.

Moreover, patterns by different age groups presented that the younger (≤29 years old) and older age groups (≥50 years old) have high correlations of Internet searches for coronavirus with new COVID-19 cases. This finding demonstrated high-risk perception from those age groups, even three days prior to the increase in the number of new COVID-19 cases. High-risk perception in younger age groups might be induced by massive Internet access for acquiring information and high numbers of confirmed cases in that age group (33.24%) in South Korea *(17, 24)*. Meanwhile, perceived vulnerability might be common in older age groups since older age is one of the prominent risk factors for COVID-19 mortality *(25)*, and 98.08% of fatal cases in South Korea occurred in older adults *(17)*. Accordingly, a previous study also showed that the older age group has a higher risk perception *(7)*.

By contrast, the age group of 30 to 49 years old only showed low to moderate correlations even for three days before the event. It might be due to the lower percentage of confirmed cases (23.94%) in that age group compared with that in the younger age group (≤29 years), which could also influence the health risk perception. Meanwhile, online queries for coronavirus test showed the high-risk perception in the 35–39-years age group. These findings illustrate that adults perceived the coronavirus test-related information as more important than disease-related knowledge. It might be also influenced by massive coronavirus test conducted so far. Meanwhile, the younger (aged ≤29 years) and older age groups (aged ≥50 years old) have a different perception, making the infection-related information among the essential searches. In terms of gender, both men and women perceived coronavirus as the same level of risk, but higher in women for coronavirus test. This result is similar to that reported a previous study which showed a higher risk perception in the female group *(7)*. Hence, health risk communication should be carried in both men and women as well as in vulnerable age groups.

For device utilization, patterns demonstrated that mobile searches have greater correlations with COVID-19-related searches compared with desktop queries. High correlations for mobile searches were even observed three days prior to the disease onset. However, desktop searches showed high correlation in lag −1, which was two days late, compared with mobile searches. This finding implies that high-risk perception stimulated enormous number of mobile searches during the outbreak period. Identical results were also illustrated in previous study by Soo-Yong Shin and colleagues *(12)*. With the widespread use of mobile devices in the digital era *(20)*, this promoted a change in the behavior, from desktop to mobile device use. Therefore, the government should ensure that risk communication could be assessed easily through mobile devices.

Research findings also demonstrated that the distributions of Internet searches were higher in the location with new COVID-19 cases. This finding was similar to that in previous studies which indicated that people in affected areas have higher risk perception *(7, 11)*. However, Internet searches continued to increase in a vulnerable location such as tourist sites including Jeju. Those findings demonstrated that health risk communication is urgently needed in affected and vulnerable areas.

In brief, this study provided the depiction of community health risk perception toward COVID-19 in South Korea, which tends to be higher in the period of local and international events, along with women, certain age groups (≤29, 35–39 and ≥50-years age group), and people in the affected areas. Moreover, during the outbreak, people were more likely to access the Internet through mobile devices, which are potential channels where health risk communication can be disseminated effectively. This method demonstrated an easy and low-cost approach to estimate health risk perception during the pandemic. Since providing a rapid risk perception assessment is urgently needed in the early stage of an outbreak, combining Google and NAVER RSV could be beneficial for targeting risk communication in terms of time, population characteristics, and location. Google RSV alone only revealed the patterns according to time and location *(26)*. As online search queries might change over time, finding the best lagged time for conducting risk communication would be challenging.

## Conclusion

Community health risk perceptions toward COVID-19 outbreak in South Korea observed from Google and NAVER RSV increased during local and international events, and were higher in women, certain age groups as well as in affected areas. While NAVER RSV tends to be more sensitive against local issues, integrating Google and NAVER RSV could potentially provide varied patterns in terms of time, population characteristics, and location.

## Data Availability

Numbers of new COVID-19 cases and coronavirus tests performed on a daily basis were available on the South Korea open access data set from Kaggle by Joong Kun Lee and colleagues in collaboration with the Korea Centre for Disease Control and Prevention (KCDC) from January 20 to March 22, 2020

## Ethical approval

No need for ethical approval as used of anonymous open data.

## Conflicts of interest

None.

### Acknowledgments

We gratefully thank Joong Kun Lee and colleagues for providing open access data on COVID-19-related information in South Korea on kaggle.com.

## Author Bio

Atina Husnayain is a PhD student in Biomedical Informatics at Graduate Institute of Biomedical Informatics, Taipei Medical University. Her research interests focus on digital epidemiology, information technology-based surveillance, spatial epidemiologi, and machine learning.

## References

1. World Health Organization. WHO Director-General’s opening remarks at the media briefing on COVID-19 - 11 March 2020: World Health Organization; 2020 [updated March 11; cited 2020 March 30]. Available from: https://www.who.int/dg/speeches/detail/who-director-general-s-opening-remarks-at-the-media-briefing-on-covid-1911-march-2020.

2. World Health Organization. Coronavirus disease 2019 (COVID-19): Situation Report – 77. World Health Organization; 2020 April 6.

3. Lee J, Yu B, Kim M, Jang S, Ryoo S, Choi W, et al. Data Science for COVID-19 (DS4C). 2020.

4. Shim E, Tariq A, Choi W, Lee Y, Chowell G. Transmission potential and severity of COVID-19 in South Korea. Int J Infect Dis. 2020;93:339–44.

5. Gallo W. South Korea Tries ‘Social Distancing’ to Prevent Coronavirus Spread: VOA News; 2020 [updated March 6; cited 2020 March 30]. Available from: https://learningenglish.voanews.com/a/south-korea-tries-social-distancing-to-prevent-coronavirus-spread/5316633.html.

6. Kwon KT, Ko JH, Shin H, Sung M, Kim JY. Drive-Through Screening Center for COVID-19: a Safe and Efficient Screening System against Massive Community Outbreak. J Korean Med Sci. 2020;35(11):e123.

7. Jang WM, Kim UN, Jang DH, Jung H, Cho S, Eun SJ, et al. Influence of trust on two different risk perceptions as an affective and cognitive dimension during Middle East respiratory syndrome coronavirus (MERS-CoV) outbreak in South Korea: serial cross-sectional surveys. BMJ Open. 2020;10(3):e033026.

8. World Health Organization. Emergencies preparedness, response: What is phase 6? : World Health Organization; 2009.

9. Hubner AY, Hovick SR. Understanding Risk Information Seeking and Processing during an Infectious Disease Outbreak: The Case of Zika Virus. Risk Anal. 2020.

10. Lohiniva AL, Sane J, Sibenberg K, Puumalainen T, Salminen M. Understanding coronavirus disease (COVID-19) risk perceptions among the public to enhance risk communication efforts: a practical approach for outbreaks, Finland, February 2020. Euro Surveill. 2020;25(13).

11. Yang JZ. Whose Risk? Why Did the U.S. Public Ignore Information About the Ebola Outbreak? Risk Anal. 2019;39(8):1708–22.

12. Shin SY, Kim T, Seo DW, Sohn CH, Kim SH, Ryoo SM, et al. Correlation between National Influenza Surveillance Data and Search Queries from Mobile Devices and Desktops in South Korea. PLoS One. 2016;11(7):e0158539.

13. Zhang Y, Yakob L, Bonsall MB, Hu W. Predicting seasonal influenza epidemics using cross-hemisphere influenza surveillance data and local internet query data. Sci Rep. 2019;9(1):3262.

14. Normile D. Coronavirus cases have dropped sharply in South Korea. What’s the secret to its success? : American Association for the Advancement of Science; 2020 [updated March 17; cited 2020 March 30]. Available from: https://www.sciencemag.org/news/2020/03/coronavirus-cases-have-dropped-sharply-south-korea-whats-secret-its-success#.

15. Arin K. Still not enough face masks to go around: The Korea Herald 2020 [updated March 1; cited 2020 April 16]. Available from: http://www.koreaherald.com/view.php?ud=20200229000149.

16. OBE TG, Howard J. Masks for all? The science says yes. 2020 [updated April 13; cited 2020 April 24]. Available from: https://www.fast.ai/2020/04/13/masks-summary/.

17. Korean Center for Disease Control and Prevention. Press Release: The updates on COVID-19 in Korea as of 22 March: Korean Center for Disease Control and Prevention; 2020 [updated March 22; cited 2020 March 30]. Available from: https://www.cdc.go.kr/board/board.es?mid=a30402000000&bid=0030.

18. Paek H-J, Hove T. Risk Perceptions and Risk Characteristics. Oxford Research Encyclopedia of Communication2017.

19. World Health Organization. World Health Organization Outbreak Communication Planning Guide. Geneva: World Health Organization; 2008.

20. Liang B, Scammon DL. Incidence of online health information search: a useful proxy for public health risk perception. J Med Internet Res. 2013;15(6):e114.

21. Li C, Chen LJ, Chen X, Zhang M, Pang CP, Chen H. Retrospective analysis of the possibility of predicting the COVID-19 outbreak from Internet searches and social media data, China, 2020. Euro Surveill. 2020;25(10).

22. Kim HJ. South Korea learned its successful Covid-19 strategy from a previous coronavirus outbreak: MERS Chicago: Bulletin of the Atomic Scientists; 2020 [updated March 20; cited 2020 April 19]. Available from: https://thebulletin.org/2020/03/south-korea-learned-its-successful-covid-19-strategy-from-a-previous-coronavirus-outbreak-mers/.

23. Keller M, Blench M, Tolentino H, Freifeld CC, Mandl KD, Mawudeku A, et al. Use of unstructured event-based reports for global infectious disease surveillance. Emerg Infect Dis. 2009;15(5):689–95.

24. Korea internet and security agency. 2018 Korea internet white paper. Korea internet and security agency; 2020.

25. Zhou F, Yu T, Du R, Fan G, Liu Y, Liu Z, et al. Clinical course and risk factors for mortality of adult inpatients with COVID-19 in Wuhan, China: a retrospective cohort study. The Lancet. 2020;395(10229):1054–62.

26. Husnayain A, Fuad A, Su EC. Applications of google search trends for risk communication in infectious disease management: A case study of COVID-19 outbreak in Taiwan. Int J Infect Dis. 2020.

